# Multi-Omic Analyses Characterize the Ceramide/Sphingomyelin Pathway as a Therapeutic Target in Alzheimer’s Disease

**DOI:** 10.1101/2021.07.16.21260601

**Authors:** Priyanka Baloni, Matthias Arnold, Herman Moreno, Kwangsik Nho, Luna Buitrago, Kevin Huynh, Barbara Brauner, Gregory Louie, Alexandra Kueider-Paisley, Karsten Suhre, Andrew J. Saykin, Kim Ekroos, Peter J. Meikle, Leroy Hood, Nathan D. Price, The Alzheimer’s disease Metabolomics Consortium, P. Murali Doraiswamy, Cory C. Funk, Gabi Kastenmüller, Rebecca Baillie, Xianlin Han, Rima Kaddurah-Daouk

## Abstract

Dysregulation of sphingomyelin (SM) and ceramide metabolism have been implicated in Alzheimer’s Disease (AD). Genome-wide and transcriptome wide association studies have identified various genes and genetic variants in lipid metabolism that are associated with AD. However, the molecular mechanisms of sphingomyelin and ceramide disruption remain to be determined. Evaluation of peripheral lipidomic profiles is useful in providing perspective on metabolic dysregulation in preclinical and clinical AD states. In this study, we focused on the sphingolipid pathway and carried out multi-omic analyses to identify central and peripheral metabolic changes in AD patients and correlate them to imaging features and cognitive performance in amyloidogenic mouse models. Our multi-omic approach was based on (a) 2114 human post-mortem brain transcriptomics to identify differentially expressed genes; (b) *in silico* metabolic flux analysis on 1708 context-specific metabolic networks to identify differential reaction fluxes; (c) multimodal neuroimaging analysis on 1576 participants to associate genetic variants in SM pathway with AD pathogenesis; (d) plasma metabolomic and lipidomic analysis to identify associations of lipid species with dysregulation in AD; (e) metabolite genome-wide association studies (mGWAS) to define receptors within pathway as potential drug target. Our findings from complementary approaches suggested that depletion of S1P compensated for AD cellular pathology, likely by upregulating the SM pathway, suggesting that modulation of S1P signaling may have protective effects in AD. We tested this hypothesis in APP/PS1 mice and showed that prolonged exposure to fingolimod, an S1P signaling modulator approved for treatment of multiple sclerosis, alleviated the cognitive impairment in mice. Our multi-omic approach identified potential targets in the SM pathway and suggested modulators of S1P metabolism as possible candidates for AD treatment.

## Introduction

To date, approximately 400 trials of experimental Alzheimer’s treatments have failed^1^. In the wake of such large-scale failure, additional hypotheses have been proposed to accelerate strategies for treatment and researchers are pursuing alternative approaches, with a greater focus on the complex mechanisms underlying this neurodegenerative disease. In an effort to address this knowledge gap, the NIH-funded Accelerating Medicines Partnership - Alzheimer’s Disease (AMP-AD) has successfully generated new hypotheses and insights around Alzheimer’s disease (AD) and produced large, publicly available datasets. The knowledge gained from this initiative will lead a major paradigm shift in research focus, resulting in novel targets and testable hypotheses that are currently being investigated in clinical phase 1 and 2 trials aimed at neuroprotection and anti-neuroinflammation^2^. These new hypotheses also suggest potential drug repositioning and development.

While the central neuropathological features of AD are accumulation of misfolded β-amyloid (Aβ) plaques and phosphorylated tau proteins, brain atrophy and neuronal loss are equally important. The relationship between Aβ accumulation, tau phosphorylation and neuronal loss is unclear. What is clear is that AD etiology is multifactorial, with genetic contributions, protein mis-trafficking and turnover, altered glucose metabolism and lipid metabolism failures^3^. Recent studies have clarified the important relationship between the immune system and lipid metabolism and more than half of the genes implicated in AD via genetic association screens are linked to lipid metabolism and inflammation^4^. Exploring how these genes factor into AD pathophysiology over the last few years is starting to increase our understanding of the role of lipid metabolism in AD. APOE4, the strongest genetic risk factor for late onset AD, is centrally involved in lipid metabolism, including the transport of cholesterol to neurons from astrocytes^5^. Additionally, several independent genetic association studies have reported replicable associations of the *APOE* locus with blood levels of sphingolipid species^6,7,8^. Lipids, including SMs, have been shown to be disrupted in AD^9,10,11^. However, the impact of lipid dysregulation on AD pathogenesis are not fully understood.

Brain lipids constitute ∼50% of the brain’s dry weight with myelin, a proteolipid, composed of 70-80% lipids^12^. Several lines of supporting evidence implicate various sphingolipids in neuronal signaling and toxicity^13,14^. Sphingomyelin (SM) primarily resides in two locations within the brain: 1) Lipid rafts, found in neurons, astrocytes, and microglia where they are involved in several aspects of signal transduction and homeostasis of the brain; 2) the membranous myelin sheath that insulates many nerve cell axons^15^. As part of lipid rafts, SMs are involved in signal transduction and the regulation of inflammatory processes and response to oxidative stress^16^. Our previous studies^17–20^ indicated a complex pattern of deregulation in the sphingolipid metabolism, including ceramides, in the early stages of AD. We have also reported changes at the gene expression level of the myelin network in AD^21^. Hydrolysis of sphingomyelin produces ceramide (Cer). Ceramides are the simplest of sphingolipids, are neurotoxic and induce apoptosis^22,23^. Ceramides are known to mediate the relationship between Aβ and neurodegeneration^24^. Increasing Aβ levels elevate SM phosphodiesterase (SMase) activity leading to an increase in Cer^25,26^. It is suggested that the increase in ceramides boosts BACE-1 activity^27^, which cleaves APP in two soluble Aβ. Sphingosine-1-phosphate (S1P), is an important neuroprotective signaling molecule and product of the SM pathway that blocks SMase activity^28^ and inhibits amyloid precursor protein (APP) secretion^29^. By understanding the changes in SM/Cer homeostasis and their underlying mechanism, we can better understand how perturbations in the SM pathway contribute to neurodegeneration.

As part of normal homeostasis, microglia constantly surveil the brain parenchyma. In development, and throughout normal life-span, they remove neuronal synapses, eliminate dying neurons, and clean up myelin debris ^30–33^. Sphingolipid-rich neuronal and myelin membranes captured through these processes undergo lysosomal degradation within microglia. This degradative process is facilitated by a lipid-sensing receptor, TREM2, that is activated by various lipids (including sphingolipids, sphingomyelin, and sulfatide). TREM2-deficient microglia phagocytose myelin debris but fail to clear myelin cholesterol, resulting in cholesteryl ester (CE) accumulation. CE increase is also observed in APOE-deficient glial cells, reflecting impaired brain cholesterol transport^34^. Recent studies have begun to elucidate the important role of microglia in AD, with evidence for differences in microglial subpopulations, related to myelin clearance and activation^35–41^.

In this study, we used human *in vivo* data and post-mortem brain data to finely characterize the SM pathway for molecular links to AD pathogenesis. We identified metabolic readouts that can be utilized to link observed molecular changes back to potential intervention targets, which were experimentally validated in animal models resulting in repurposed drug for AD. We started with gene expression profiling from three large cohorts to identify differentially expressed genes in the SM pathway. We then utilized constraint-based modeling to narrow down the search space to identify potential changes in metabolic fluxes between cognitively normal and AD individuals. Next, we identified genetic variants within SM pathway and their association with changes in neuroimaging using large multicenter biomarker study. We then identified changes in plasma lipidomic species within the SM pathway with specific SNPs using data from ADNI and AIBL cohort. Using complementary approaches, we identified sphingosine 1-phosphate (S1P) regulating the balance in the pathway. We tested our hypothesis and demonstrated that fingolimod, a S1P receptor (S1PR) modulator which causes internalization of S1PR, is able to improve cognition in APP/PS1 mice. We highlight S1P as the metabolite involved in maintaining the balance in the pathway and identifying drugs regulating S1P levels that can be repurposed for AD.

## Results

### Global transcriptomic dysregulation of the SM pathway in AD

We analyzed gene expression changes of well-characterized enzymes in the sphingolipid pathway from post-mortem brain RNA-seq data generated on seven brain regions (cerebellum, temporal cortex, dorsolateral prefrontal cortex, parahippocampal gyrus, frontal pole, inferior frontal gyrus and superior temporal gyrus) in three independent cohorts (ROS/MAP, Mayo and Mount Sinai), of 2114 brain samples as well as the cross-region, cross-study meta-analysis^42^. For this study, we manually curated the sphingolipid subsystem definition of the human genome-scale metabolic reconstruction ^43^ resulting in the identification of a set of 35 enzymes catalyzing 18 enzymatic reactions within the SM pathway (Figure 1, Supplementary Table 1). The reactions cover Cer and SM biosynthesis, as well as four exit routes (through sphinganine-1-phosphate, ceramide-1-phosphate, sphingomyelin, glycosphingolipids and sphingosine-1-phosphate). Gene expression data was available for 31 of the 35 genes, the exceptions being *CERS3, ACER1, ASAH2* and *ENPP7*. Low and/or no expression of these genes in the brain was confirmed in the GTEx Portal.

**Figure 1:**
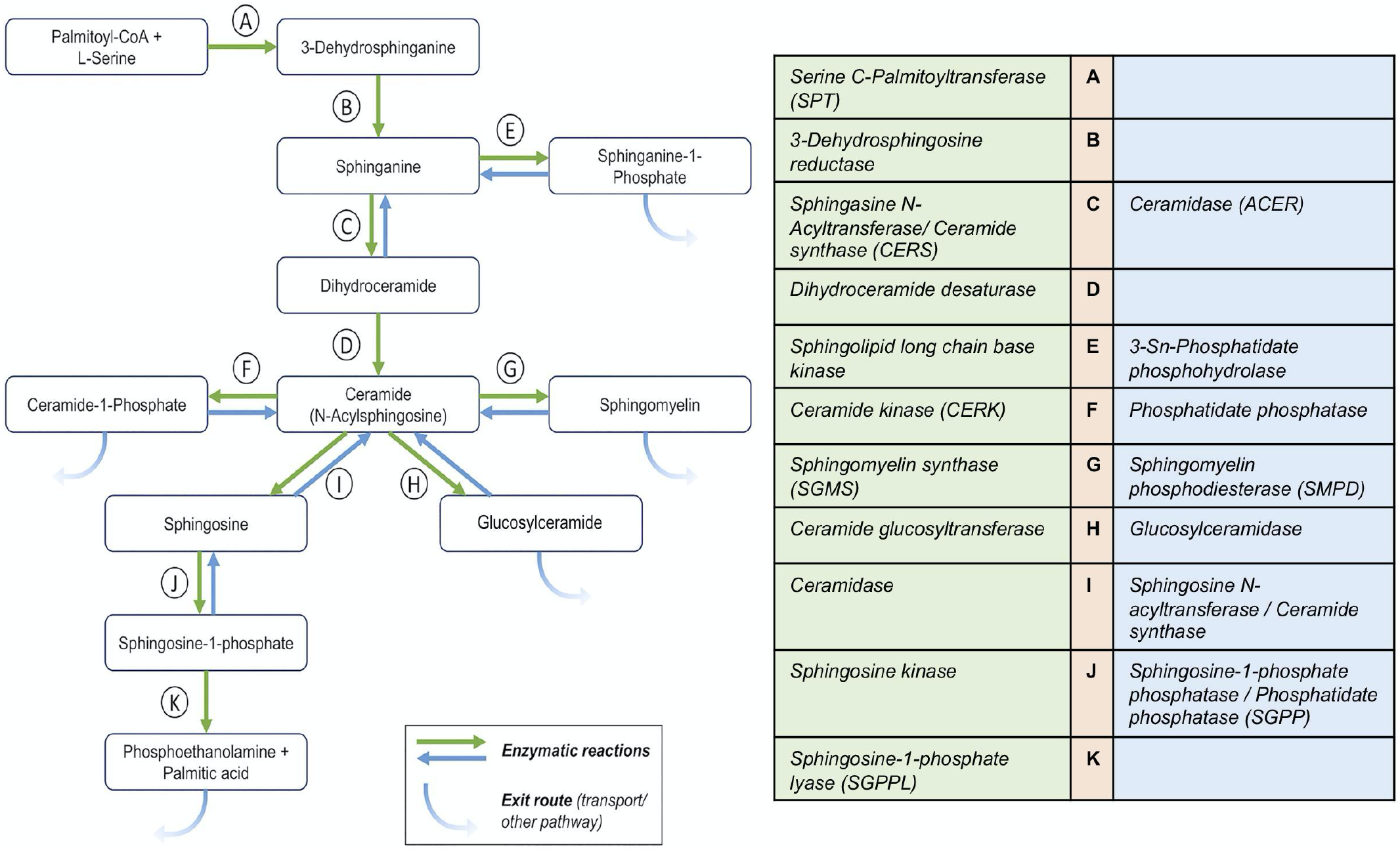
Overview of sphingolipid pathway manually curated from the Recon3D model. The metabolites participating in reactions are represented in boxes. The arrows for reactions A-K are colored based on the direction in the pathway. Some reactions are not reversible (single arrows). The table on the right lists the catalyzing enzymes in the sphingolipid pathway in humans and are denoted with the same color code as the reaction arrow.

Analysis of differential gene expression showed significant (FDR-corrected) gene expression changes in brain tissue of AD cases vs. controls for 20 of the genes (Supplementary Table 2). Of those 20, 19 showed differential expression in one or more studies/brain regions. Fourteen of these were also detected in the meta-analysis. Transcripts of *SPTLC3* were not measured in all brain regions, hence it was not reported in the meta-analysis. *DEGS1*, on the other hand, was insignificantly but consistently upregulated in the single studies, leading to a detectable significant overall upregulation in the meta-analysis. Almost all of the genes showed significantly higher expression in AD cases, consistent across all brain regions. Exceptions were *CERS5* (lower levels in cerebellum of AD cases; not significant in the meta-analysis), *CERS6* (higher levels in cerebellum vs. lower levels in the parahippocampal gyrus of AD cases; not significant in the meta-analysis), and *SMPD3* (lower levels in temporal cortex of AD cases; also significant in the meta-analysis).

### Differential reaction fluxes for SM and Cer associated reactions

We used brain region-specific metabolic reconstructions^44^ and integrated the post-mortem brain RNA-seq data with them to identify reactions that had differential fluxes in AD vs cognitively normal (CN) or control individuals. For the dorsolateral prefrontal cortex, we identified reactions catalyzed by serine palmitoyltransferase (SPT, encoded by *SPTLC1/2/3*, enzyme *A in Figure 1*), sphingomyelin synthase (SMS, encoded by *SGMS1/2*, enzyme *G in Figure 1)*, and ceramide kinase (CERK, encoded by *CERK*, enzyme *F in Figure 1*) as having significant flux differences as shown in Figure 2. SPT catalyzes the first step in the biosynthesis of sphingolipids condensing serine and palmitoyl-CoA to form 3-ketosphinganine, which is the rate-limiting step in the synthesis of SMs (Figure 1). For this reaction, we found significant differences in flux values comparing AD and mild cognitive impairment (MCI) cases (Figure 2A). Sphingomyelin synthase synthesizes sphingomyelin from ceramide. Here, we observed AD transcriptomes having higher reaction fluxes compared to the CN samples (Figure 2B). We further identified flux differences for the reaction catalyzed by ceramide kinase (phosphorylation of ceramide to form ceramide-1-phosphate) in AD and CN samples (Figure 2C).

**Figure 2:**
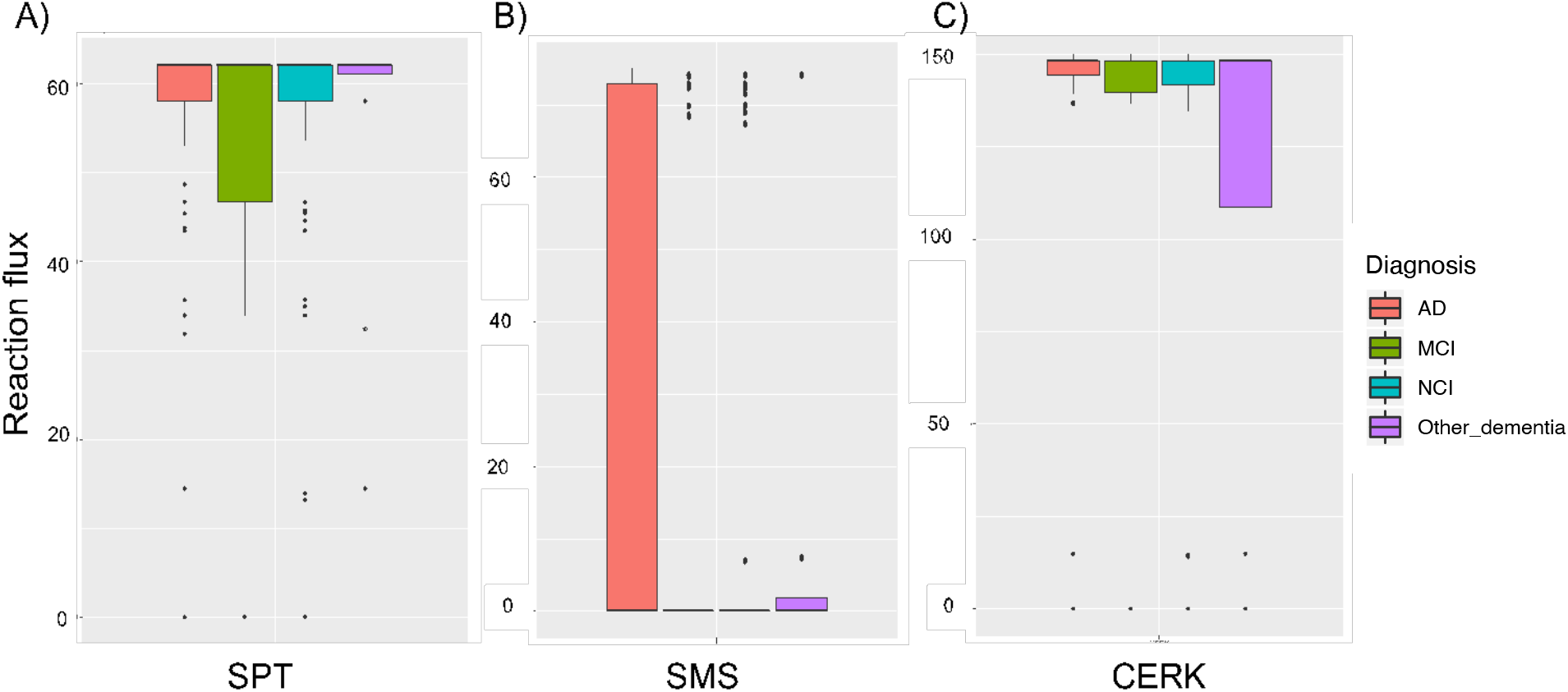
Box plot of reaction fluxes for (a) serine palmitoyl transferase, (b) sphingomyelin synthase, and (c) ceramide kinase reactions. The orange, green, blue and purple bars correspond to AD, MCI, NCI and other dementia.

### Genetic association studies and multimodal neuroimaging analysis link SM pathway to AD pathogenesis

Using gene-based association analysis in 1,576 participants of the AD Neuroimaging Initiative (ADNI) phases 1, GO and 2, we identified genetic variants in the coding regions linked to seven of the 35 genes in the SM pathway to be significantly associated with AD and its (bio)markers, which covered the whole spectrum of Amyloid, Tau, Neurodegeneration, Cognition (A-T-N-C) measures^45^ (Supplementary Table 3). A-T-N-C measures of AD are calculated by investigating genetic associations of CSF biomarker levels, brain atrophy (magnetic resonance imaging), brain glucose metabolism ([^18^F]FDG PET), cognition, and clinical diagnosis. In this analysis, Bonferroni-significance was determined by gene-specific thresholds correcting for the number of all genetic variants assigned to a certain gene. Associated markers included CSF Aβ_1-42_ (*CERS2*, enzyme *C in Figure 1*), the ratio between CSF tau (both total tau and p-tau) and CSF Aβ_1-42_ (*ACER2 (*enzyme *C in Figure 1), PLPP2*), region of interest-based measures of [^18^F] fluorodeoxyglucose positron emission tomography (FDG-PET; *CERS3, SPHK2*), cognitive performance measured, among other, by the 13-item cognitive subscale of the AD assessment scale (ADAS-Cog.13; *CERS6, DEGS1*), and clinical AD (*CERS3, CERS6, DEGS1*). Furthermore, a detailed whole brain analysis of brain glucose metabolism (FDG PET) on a voxel-wise levels showed that rs1847325 in *CERS3* (enzyme C in Figure 1) and rs281380 in *SPHK2* (J in Figure 1) were significantly associated with increased brain glucose metabolism in the bilateral frontal, parietal, and temporal lobes (colored regions with corrected p-value < 0.05; Supplementary Figure 1). Previously, a study on clinico-pathologic AD dementia^46^ yielded an association with *SMPD2* (enzyme G in Figure 1) that is Bonferroni-significant at the gene-wide level.

A less stringent p-value cutoff (adjusting for multiple testing by permutation as SNPs are correlated due to linkage disequilibrium) identified variants in two additional genes, *SPTLC3* (enzyme A in Figure 1) and *SGMS1* (enzyme G in Figure 1). *SPTLC3* was associated with cognitive performance (corrected p-value = 0.02; Figure 3A), brain atrophy in focal regions of the bilateral temporal and frontal lobes (determined by detailed surface-based whole-brain analysis of cortical thickness measured from MRI scans on a vertex-wise level; colored regions with corrected p-value < 0.05; Figure 3B) and FDG-PET measures in the bilateral temporal and parietal lobes (colored regions with corrected p-value < 0.05; Figure 3C). *SGMS1* was associated with brain glucose metabolism measured by region of interest-based FDG-PET (corrected p-value = 0.02; Figure 3D) that was mapped by whole brain analysis to the bilateral temporal, parietal, and frontal lobes, as well as the hippocampus (colored regions with corrected p-value < 0.05; Figure 3F). In addition, surface-based whole brain association analysis showed a significant association with cortical thickness in the bilateral temporal, parietal, and frontal lobes, with the strongest association located in the entorhinal cortex (colored regions with corrected p-value < 0.05; Figure 3E).

**Figure 3:**
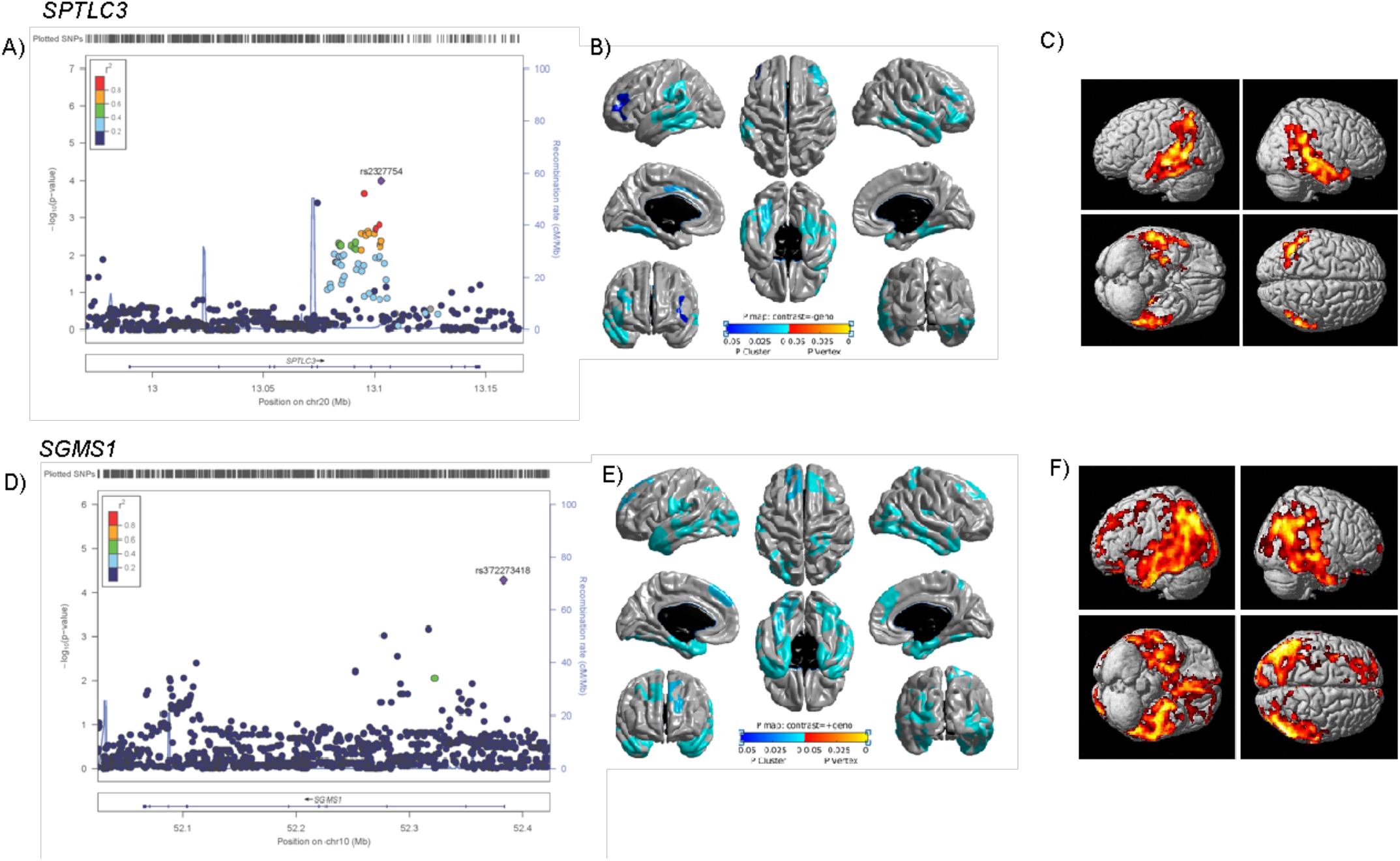
Association of genetic variants in *SPTLC3* and *SGMS1* with structural (MRI) and molecular (FDG-PET) neuroimaging phenotypes. A) Gene-based association analysis of *SPTLC3* with cognitive performance (Rey auditory verbal learning test total score). D) Gene-based association analysis of *SGMS1* with global brain glucose metabolism. B & E) Surface-based whole brain analysis of cortical thickness (brain atrophy measured from MRI scans) for *SPTLC3* and *SGMS1*. C & F) Voxel-based whole brain analysis of brain glucose metabolism measured from FDG PET scans for *SPTLC3* and *SGMS1*.

### SM (d34:1)/SM (d43:1) ratio as a strong intermediate trait for sphingolipid dysregulation in AD

Sphingomyelin species (SMs) of differing lengths have been implicated in early vs. late stages of AD^19^. SM (d34:1) is associated with CSF A β 1-42 pathology, while SMs with longer fatty acid chains (≥C20) are correlated with brain atrophy and cognitive decline. Utilizing the concept of metabolite ratios^47^, which enables both removal of potentially remaining technical variance and modeling of enzymatic/pathway activity^8^, we selectively screened ratios of shorter chain SMs (<C20) and longer chain SMs (≥ C20) in the ADNI-1 dataset (n = 732) similar to Toledo et al.^19^. This revealed the ratio of SM (d34:1) and SM (d43:1) as the metabolic trait most significantly associated with diagnosis of clinical AD (p-value = 1.70 × 10-4, P_gain_ = 178.37), brain atrophy in regions implicated in AD^48^ (p-value = 7.64 × 10-6, P_gain_ = 687.57) as well as cognition (measured by ADAS-Cog. 13; p-value = 4.36 × 10-6, P_gain_ = 2544.51). The modified Alzheimer’s Disease Assessment Scale cognitive subscale (ADAS-Cog 13-item scale)^49^ has all the original ADAS-Cog items with additional items that were aimed to increase the number of cognitive domains and range of symptom severity.

To expand upon and further validate this finding, we examined the same cohort (ADNI1) using a more comprehensive lipidomics method covering a broader range of sphingolipids. In total, 112 sphingolipids were examined in serum samples (n = 754), where chromatography enabled separation of some isomeric and isobaric species. Regression analysis (adjusting for age, sex, BMI, HDL-C, total cholesterol, triglycerides, APOE e4 and fasting status) between individual lipid species and lipid ratios (112 individual species, totaling 12,544 ratios) with ADAS-Cog 13 identified 3385 ratios associated with an uncorrected p-value of < 0.05 and 1552 significant post FDR correction (Supplementary Table 4). This analysis confirmed that ratios of short to longer chain sphingomyelins, in particular the ratio of SM(d34:1)/SM(d43:1), presented with a positive association with ADAS-Cog 13 scores (FDR corrected p-value of 3.98 × 10^−2^).

### Using the SM species for genetic screening and pathological markers in AD

To link SM readouts associated with AD to genes, we performed metabolite genome-wide association studies (mGWAS) with the three SMs reported to be associated with markers of AD in Toledo et al., as well as the selected ratio of SM(d34:1)/SM(d43:1). The discovery analysis was performed in a subset of 674 ADNI-1 participants from Toledo et al.^19^ that had genome-wide genotyping data available. While the three single SM species did not yield significant results, the SM ratio was associated with *SPTLC3 (*enzyme *A in Figure 1)* at genome-wide significance corrected for 4 metabolic traits (lead SNP rs680379, p-value = 1.01 × 10^−9^). This association replicated a previous finding in a larger population-based mGWAS investigating metabolite ratios (rs168622, *r*^*2*^ = 0.98 with rs680379, p-value = 5.2 × 10^−25^)^20^.

Lookup of the *SPTLC3 (*enzyme *A in Figure 1)* locus using the large collection of metabolite-genotype associations in the *SNiPA* database^50^) revealed significant links to several additional SM species. To obtain a comprehensive map of genetic influences on SM levels across the whole SM pathway, we again used gene-based association analyses including all 35 genes in the pathway analogously to the analysis of associations with markers of AD. To this end, we used an expanded set of 1,407 ADNI participants with SM readouts and genome-wide genotype information available, as well as two large population-based mGWAS studies that included SM levels^6,8^. We found genome-wide and gene-wide significant associations with a set of 14 related SMs for six genes (Figure 4, Supplementary Table 5). Three of the encoded enzymes are involved in SM synthesis (*SPTLC3, CERS2, CERS4*), while the other three function in synthesis and degradation of S1P (*SPHK2, SGPP1, SGPL1*), a central exit route of the pathway. Notably, the significant associations include all three SMs identified by Toledo et al.^19^ (SM (d33:0), SM (d34:1), and SM (d38:2)), highlighting a potential role for S1P metabolism and signaling in AD pathogenesis.

**Figure 4:**
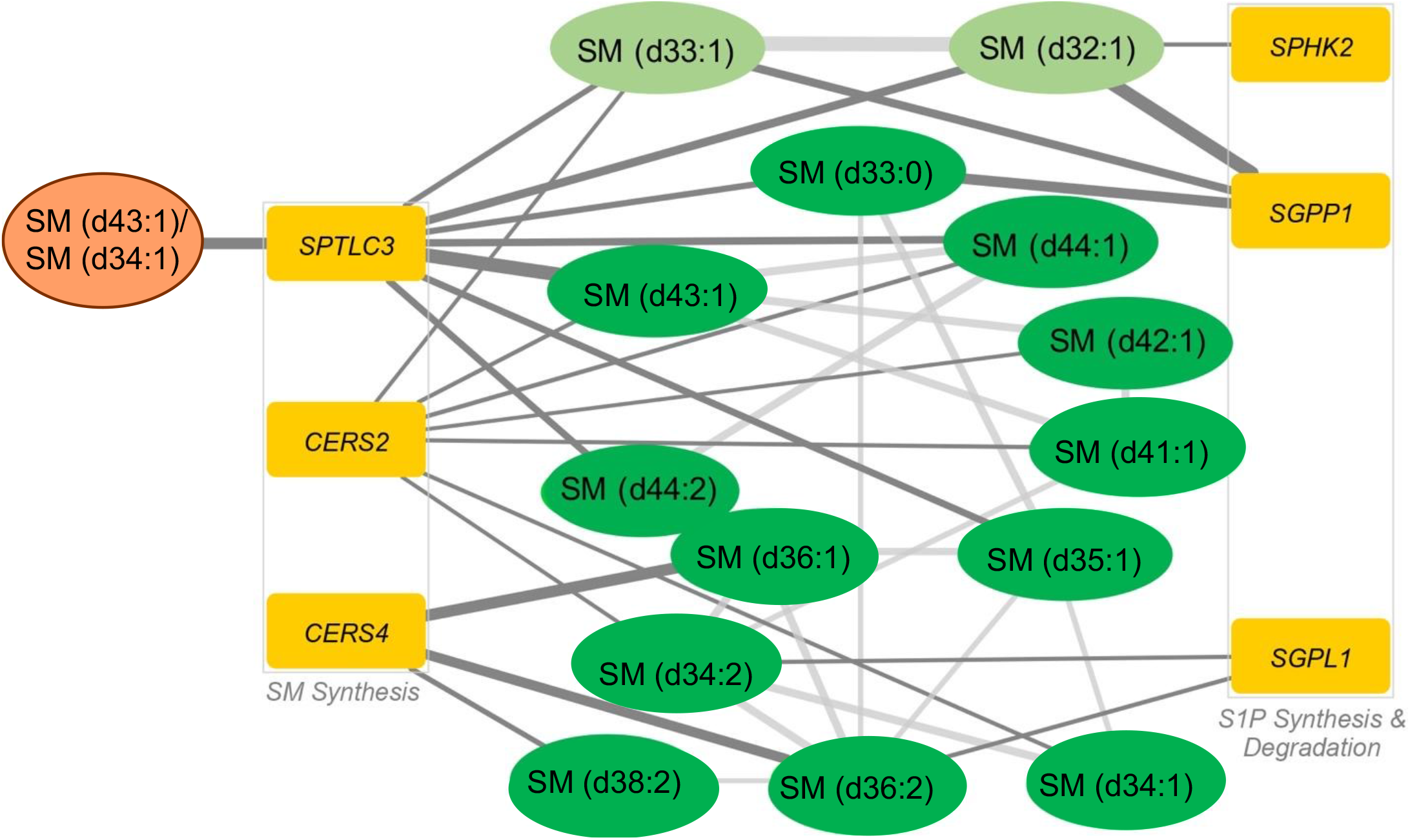
Hybrid network of genetic associations revealed by gene-based association studies and significant partial correlations of detected sphingomyelins^8,19^. The six identified genes can be grouped into two categories: global sphingomyelin synthesis and synthesis and degradation of sphingosine-1-phosphate. The selected SM ratio is colored in orange, other SM species are in green (light green: non-targeted metabolomics in Shin et al.^8^; dark green: targeted metabolomics in ADNI and Draisma et al.^6^), and genes are in dark yellow.

### Treatment of amyloidogenic APP/PS1 mice with Fingolimod

To functionally investigate involvement of deregulated S1P metabolism in amyloid pathology along with strategies to counter potentially links to AD pathogenesis, we applied a drug repositioning approach by treating amyloidogenic APP/PS1 mice with fingolimod (FTY720), an FDA-approved drug for the use in the relapsing-remitting form of multiple sclerosis^51^. The APP/PS1 mice are transgenic mice expressing chimeric amyloid precursor protein (APP) and mutant human presenilin 1 (PS1) and are valuable models to study AD progression and effect of drugs on AD^52^. The immunomodulating compound is a sphingosine analog that, after endogenous phosphorylation by sphingosine kinases 1 and 2, broadly binds to S1P receptors (S1PR1/3/4/5)^53,54^.

To corroborate previously published data with similar ages, we phenotyped APP/PS1 (n = 6, 50% female) and WT mice (n = 6, 50% female) at 7 months old (m.o.) using a battery of behavioral tests. Novel object recognition (NOR) test and Barnes Maze task were utilized to assess episodic and spatial memory respectively. After behavioral testing, we evaluated synaptic transmission using electrophysiology experiments at the Schaffer collateral-CA1 synapse. APP/PS1 mice had a significant deficit in the NOR test (Discrimination index (DI)=-0.18 ± 0.18, Figure 5A) compared to WT mice (DI = 0.33 ± 0.24; t_5_ = 3.25; p-value = 0.02). During the Barnes Maze task APP/PS1 mice showed a mild deficit in spatial learning abilities (t_36_ = 3.098; p-value = 7.5 × 10^−3^ on 1^st^ day and t_36_ = 3.156; p-value = 7.5 × 10^−3^ on 2^nd^ day) (Figure 5B) and memory retention showed by the less time spent in the target quadrant (30.39 ± 4.8 vs. 57.97 ± 4.4, t_8_ = 3.939; p-value = 4.3 × 10^−3^) (Figure 5C) compared to their WT littermates. Furthermore, they showed abnormal long term potentiation (LTP) both at the early and maintenance phase in the CA3-CA1 synapse (Figure 5D,E,F), where APP/PS1 mice could not maintain the potentiation 40 min after HFS (112.7% ± 1.8) compared to WT mice (188.4% ± 3.06; F_(1_,_38)_ = 95.37, p-value < 0.0001). Of note, basal synaptic transmission as evaluated by the input-output relationship was similar in the two groups (data not shown).

**Figure 5.**
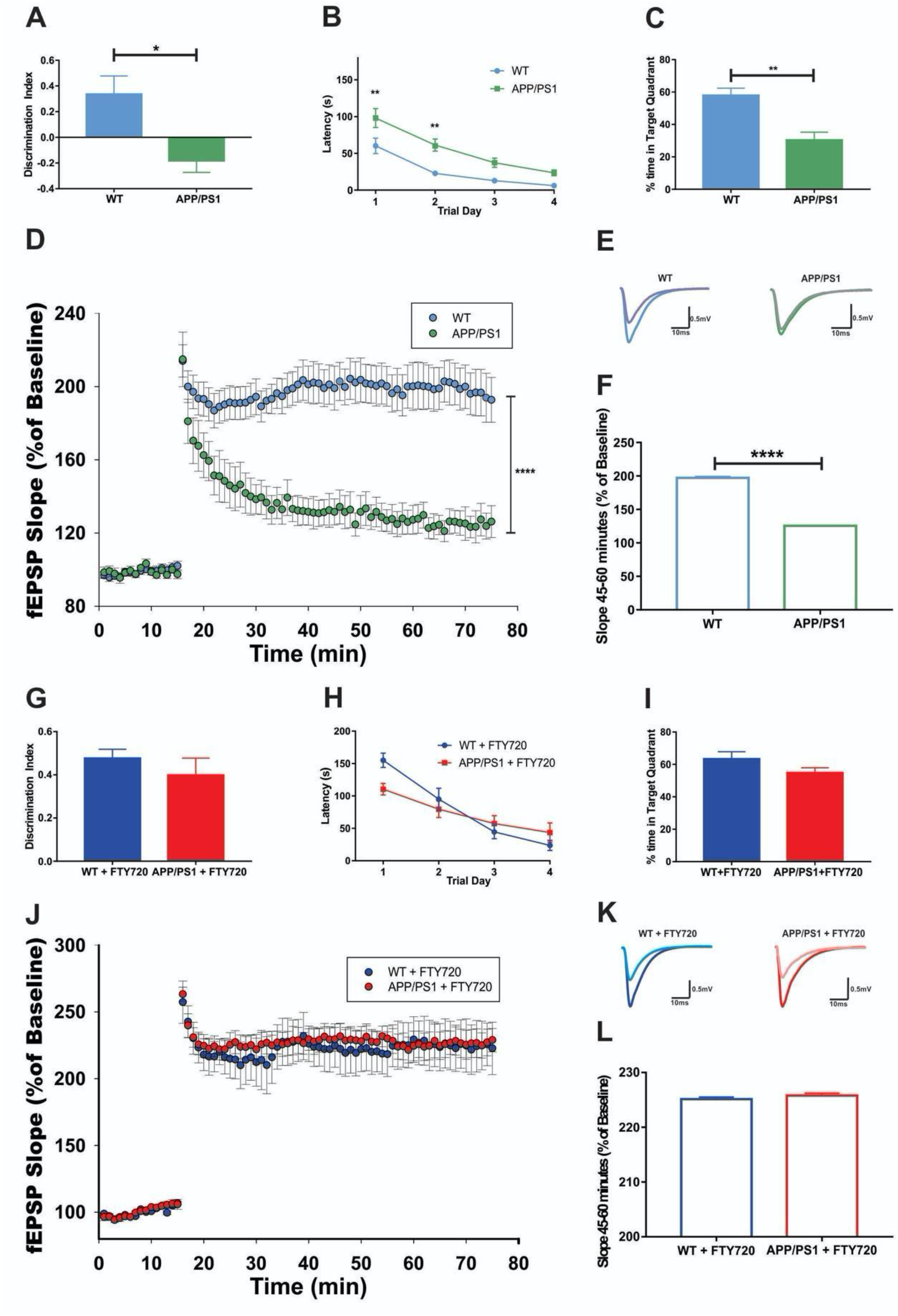
APP/PS1 mice hippocampal dependent behavior and synaptic transmission assessment and effect of fingolimod treatment. **A**. Exploration time spent on the novel object in the NOR test session. Data is expressed as a discrimination index ± SEM. **B**. Barnes maze performance during training days. Acquisition learning trials were performed, and the time it took to locate and enter into the escape box was reported in seconds. The average performance of four trials per day was expressed as mean ± SEM. A shorter latency indicates faster spatial learning. **C**. Probe trial was performed on day 5 of the Barnes Maze protocol, during which the escape box was removed. The percentage of time spent inside the target quadrant (the previous escape box location) is plotted ± SEM. A larger percentage of time indicates better spatial memory. **D**. LTP timeline. Plotted are normalized evoked excitatory post-synaptic potentials (EPSPs) slopes (Y) vs. recording time (X). The first 20 min of evoked responses were normalized and used as the baseline responses of LTP. **E**. Representative analog traces of evoked EPSPs before (light purple and grey) and after (blue and green) high frequency stimulation (HFS). **F**. The magnitude of LTP was determined according to the responses between 45 and 60 min after the HFS. Data represent mean fEPSP Slope ± SEM. (*n* = 6 mice in each group). **G**. APP/PS1 mice treated with fingolimod were tested in NOR task. Data is expressed as a discrimination index ± SEM. Fingolimod treatment significantly enhance the discrimination index of the APP/PS1 mice. **H**. Average performance of four trials per day expressed as mean ± SEM and **I**. Percentage of time spent inside the target quadrant ± SEM at the Barnes Maze task of APP/PS1 fingolimod treated mice. Fingolimod mitigated the spatial learning deficits of the APP/PS1 at 9 mo. **J**. LTP of the CA3 to CA1 synapse timeline. **K**. Representative analog traces of evoked EPSPs before (light blue and light red) and after (blue and red) HFS. **L**. The APP/PS1 treated mice group significantly augmented the normalized slope of fEPSP after HFS.

In the second experiment, mice were treated with fingolimod (1mg/Kg/day) for eight weeks (APP/PS1, n = 6, 50% female; WT, n = 6, 50% female; all mice 7 m.o. at the beginning of treatment). We found that fingolimod-treated APP/PS1 mice had similar values in NOR-DI as compared to treated WT mice (0.40 ± 0.13 vs. 0.47 ± 0.08; t_6_ = 0.95; p-value = 0.38) (Figure 5G). Comparison of APP/PS1 treated vs. WT treated mice shows that the percentage of time spent in the target quadrant for treated APP/PS1 was similar to treated WT mice (54.81 ± 9.15 vs. 63.46 ± 11.51, t_14_ = 1.677; p-value = 0.12) (Figure 5I). Latency across training days also showed no difference between groups (t_3_ = 0.4697; p-value = 0.67) (Figure 5H). Remarkably, fingolimod treatment of APP/PS1 mice significantly augmented the normalized slope of fEPSP at the CA3 to CA1 synapse after HFS (Figure 5J,K). Comparison of the average slope percent change during the last 20 min of the maintenance phase also showed no difference (225.8% ± 1.86 vs. 225.1% ± 1.78; t_38_ = 1.163; p = 0.25) (Figure 5L).

We then compared APP/PS1 fingolimod treated vs. untreated APP/PS1 (9 m.o. 50% female, n=6 treated, n=4 untreated), and analysis of the latency during training showed a deficit in untreated compared to treated APP/PS1 mice (RM-ANOVA F = 3.1; p = 0.041) (Supplementary Figure 2). These data indicate that prolonged S1P pathway modulation can rescue both the proposed cellular mechanism of hippocampus related memory (synaptic LTP) and the cognitive deficits per se in amyloidogenic APP/PS1 mice.

## Discussion

In this study, we systematically analyzed the SM pathway for multi-omics links to pathogenic processes in AD. An overview of this study is shown in Figure 6. We were able to replicate the findings in human post-mortem samples, *in vivo* samples and mouse models. The key findings from the multi-omics work are: a) Using post-mortem brain transcriptome data of 2114 samples, we identified differentially expressed genes in the SM pathway of AD patients; b) comparison of 1708 context-specific metabolic reconstruction of the brain regions showed differences in the reaction fluxes for AD and CN samples; c) multimodal neuroimaging analysis of 1576 individuals identified genetic variants linked to genes in SM pathway and associated with AD pathogenesis; d) plasma metabolomic and lipidomic analysis identified the SM(d43:1)/SM(d34:1) ratio as a strong intermediate trait for sphingolipid dysregulation in AD; e) metabolite genome-wide association studies (mGWAS) identified S1P metabolite as potential AD drug target; and f) experimental analyses of amyloidogenic APP/PS1 mice treated with fingolimod revealed beneficial effects of S1P modulation and alleviated the cognitive impairment in mice.

**Figure 6:**
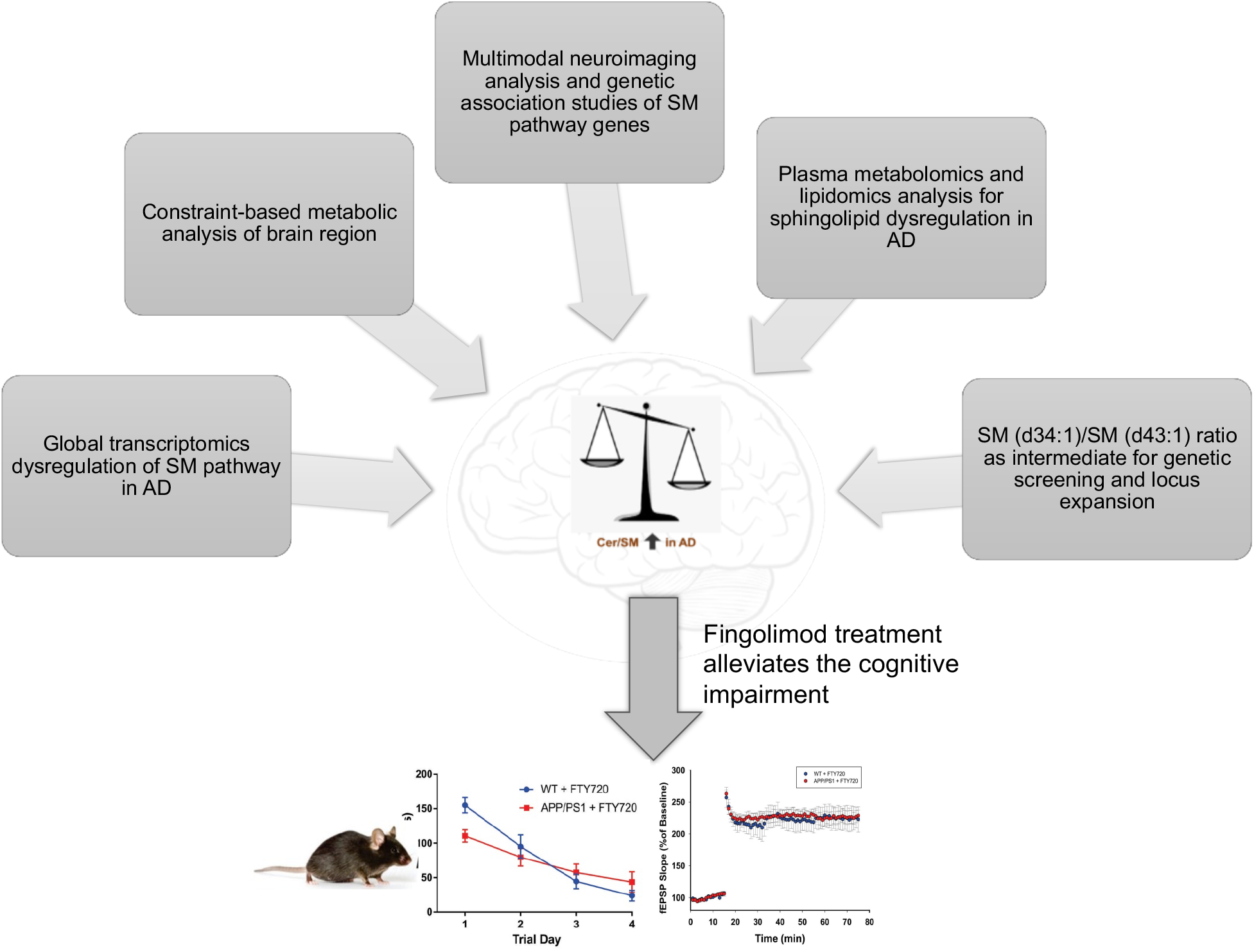
Overview of the study. We used information obtained from (a) post-mortem brain transcriptomics analysis; (b) metabolic networks of brain regions; (c) genetic variants associated with AD biomarkers including neuroimaging endophenotypes (MRI and FDG PET); (d) plasma metabolomics and lipidomics analysis; (e) genetic screening using SM (d43:1)/SM (d34:1) ratio. The balance of ceramide and sphingomyelin levels in AD could be maintained by modulating S1PR activity. The hypothesis was tested on APP/PS1 mice treated with fingolimod.

We demonstrated that, on the gene expression level, the SM pathway is globally dysregulated across brain regions in samples of AD cases compared to controls. We found that 20 out of 35 genes encoding the core enzymes in the pathway are significantly differentially expressed in the AD population. The only sub-pathway that appears to be unaffected by or uninvolved in the disease is the synthesis and recycling of glycosphingolipids. Using constraint-based metabolic networks of brain regions integrated with post-mortem brain transcriptome data, we further show that the differential expression of the enzymes involved in at least three reactions are predicted to result in significant flux differences in AD cases versus controls. While flux differences cannot be directly interpreted with respect to the resulting metabolic changes, there is ample evidence from metabolomics studies that the pathway exhibits differential output in AD.

We next assessed the association of genes in the SM pathway with A-T-N-C measures of AD by investigating genetic associations of CSF biomarker levels, brain atrophy (magnetic resonance imaging), brain glucose metabolism ([^18^F] FDG PET), cognition, and clinical diagnosis. Ten of the 35 genes in the pathway showed significant associations with at least one (endo)phenotype at the gene level. Although not genome-wide significant, this large coverage of genes in the SM pathway suggests that there might be at least a small fraction of genetic risk predisposition to AD attributable to the pathway as a whole. Using SM levels as intermediate traits for genetic association, screening further revealed six central enzymes in the pathway to be genetically influencing levels of a network of 14 SM species. As all of the genetic variants associated with SM levels were linked to the respective enzymes via expression quantitative trait loci, this indicates that some of the genetic links between the pathway and markers of AD may be mediated by altered regulation of SM levels via genetically influenced differential gene regulation.

While associations from the analysis of differential gene expression in brain tissue as well as from the phenotype GWASs were broad and generally implicated SM pathway function, the associations from the SM mGWASs linked two central pathway routes: global SM synthesis and S1P metabolism. Based on previous mGWAS analysis, genetic associations with core enzymes involved in the primary synthesis of SM metabolites are expected. However, the specific association with one particular exit route out of the pathway (via sphingosine and S1P) is striking. Five of the six detected genes (*SPTLC3, CERS2, CERS4, SPHK2* and *SGPL1*) were also found to be significantly linked to AD either through differential gene expression or via genetic associations or both, which suggests that S1P metabolism may be relevant to disease.

S1P is known to be involved in endothelial barrier function in a context-dependent manner. Decreased S1P by lipopolysaccharide (LPS) treatment produced blood brain barrier (BBB) abnormalities, and increased activity of SGPP1 and S1PR^28^. Chronic BBB leakiness is associated with cognitive impairment, but not with signs of brain inflammation^29^. S1P in general increases neuronal and circuit excitability^30,31^. Depletion of the S1P producing enzyme SphK1 induces an impairment of mossy fiber—CA3 LTP and deficits in spatial reference memory^32^. Depletion of SphK2 produced lower levels of hippocampal S1P, reduced histone acetylation and deficits in spatial memory as well as impaired contextual fear extinction^33^. Thus, S1P, SphK1 and SphK2 play specific roles in brain areas serving specific memory functions through intracellular S1P effects as well as signaling pathways downstream of S1P GPCRs. A recent study showed that Aβ1–42 enhanced SphK1 expression and activity after 24 h, but down-regulated them after 96 h and had no effect on Sphk2. Aβ1–42 and SKI II induced free radical formation, disturbed the balance between pro- and anti-apoptotic proteins and evoked cell death in PC12 cells while SP1 rescued part of this damage^37^. S1P may act as a second messenger, but it can also be transported to the extracellular space and may affect cell function *via* stimulation of the receptors (S1PR1–5). Two modulators of SP1R1 (Fingolimod and SEW2871) have been shown to improve Aβ mediated behavior abnormalities and decrease tau phosphorylation.

To explore the effect of drugs, we used APP/PS1 mice. Fingolimod is a sphingosine-1-phosphate receptor modulator approved for treatment of multiple sclerosis in the US. In absence of the drug treatment, we observed that APP/PS1 mice had a significant deficit in the novel object recognition test (NOR) and other array of tests as compared to the WT mice. Upon administration of fingolimod we found that fingolimod-treated APP/PS1 mice had similar values in NOR-DI as compared to treated WT mice. These results suggested that fingolimod modulated S1P pathway and was able to alleviate cognitive deficits in APP/PS1 mice.

The approach used here opens the possibility of repurposing fingolimod, or other S1P modulators, for treatment of AD. Fingolimod has been shown to modulate both amyloid and tau pathology in AD models^55^,^56^,^57^ and it has been proposed to be neuroprotective by modulating S1P signaling in the brain^58^. A recent study used network pharmacology methods and showed the probable pharmacological mechanism of fingolimod in the frontal cortex region of AD patients^59^. Rescuing both memory (synaptic LTP) and the behavior itself (substrate and end-result) with fingolimod is a compelling finding, which provides evidence for dysregulated S1P signaling in AD mice and further supports the identification of this pathway as a high priority candidate AD drug target. The effect of fingolimod in APP/PS1 on behavior and synaptic transmission can be direct or through the activation of S1P receptors or both, since they are not mutually exclusive. We feel fingolimod is likely one of several compounds approved or being tested for other neurodegenerative diseases that can be repurposed. This study integrates diverse types of multi-omics data from AD patients and an animal model to identify multiple, dysregulated steps in SM metabolism. It provides a link between SM dysregulation and changes in brain function. Furthermore, this study suggests that repurposing drugs that target SM metabolic enzymes, such as the S1P receptor, could correct the dysregulation and potentially improve memory and synaptic function.

Thus, using a multi-omics approach to analyze the big data led to the understanding of the sphingolipid pathway and strategies for novel drug discovery in AD.

## Methods

### Identification of differential gene expression in brain tissue RNA-seq data

We used the reprocessed AMP-AD RNA-seq data available from three studies – the Religious Order Study and the Rush Memory and Aging Project (ROS/MAP), the Mount Sinai Brain Bank cohort (MSBB), and the Mayo clinic RNA-seq study^42^ – covering 7 brain regions (cerebellum, temporal cortex, dorsolateral prefrontal cortex, parahippocampal gyrus, frontal pole, inferior frontal gyrus and superior temporal gyrus), as well as a published meta-analysis of these datasets42 to identify genes in the SM pathway that are differentially expressed in AD cases compared to controls. Gene expression changes were considered significant at an FDR-corrected p-value ≤ 0.05. All datasets are publicly available, see the Data Availability Statement.

### Metabolic networks of brain regions

Genome-scale metabolic networks of brain regions were reconstructed in our previous study^60^ and we used these metabolic networks for our present work. We integrated the post-mortem brain transcriptome data as mentioned in^60^. Using iMAT algorithm^61^, we generated context-specific personalized metabolic networks for each post-mortem sample in the dataset. Human cells in general do not proliferate rapidly and they tend to maintain their metabolic functions^62^. We therefore chose the biomass maintenance reaction, glutamate and glutamine exchange as the objective function for the brain regions. We used the dorsolateral prefrontal cortex samples for the present analysis. We performed flux variability analysis (FVA) to evaluate minimum and maximum flux for each reaction in the metabolic networks. If the minimum and maximum flux was 0 then the reactions were considered to be non-active and were assigned a state of 0, while the remaining reactions were considered to be active and assigned a state of 1. We carried out the analysis for all context-specific metabolic networks. We generated a matrix of binary values for all reactions in the context-specific metabolic networks. We selected the reactions that were part of sphingolipid metabolism using the subsystem definition. We used Fisher’s exact test on the binarized values of reactions to identify reactions with p-value of < 0.05 in AD versus CN samples. These reactions were identified as significant reactions in the groups. We used COBRA toolbox v3.0^63^ for metabolic analysis that was implemented in MATLAB R2018a and academic licenses of Gurobi optimizer v7.5 and IBM CPLEX v12.7.1 were used to solve LP and MILP problems.

### Neuroimaging processing and analysis

Participants of the Alzheimer’s Disease Neuroimaging Initiative (ADNI) were used in the analysis. Demographic information, imaging scan data, neuropsychological test scores, and clinical information were downloaded from the ADNI data repository (www.loni.usc.edu). As described in detail in previous studies^64,65^, T1-weighted structural magnetic resonance imaging (MRI) scans were processed by using a widely employed automated MRI analysis technique (FreeSurfer) to extract cortical thickness. Pre-processed [^18^F] FDG positron emission tomography (PET) scans were downloaded. Methods for acquisition and processing of PET scans were described previously (refs). [^18^F] FDG PET scans were intensity-normalized using a pons region of interest to create standardized uptake value ratio (SUVR) images. For surface-based whole brain analysis of cortical thickness on a vertex-by-vertex basis, the SurfStat software package (www.math.mcgill.ca/keith/surfstat/) was used to perform a multivariable analysis of generalized linear regression to examine the association of genetic variation on brain structural changes. Age, sex, years of education, intracranial volume, and magnetic field strength were used as covariates. In order to adjust for multiple comparisons, the random field theory correction method was used with p < 0.05 adjusted as the level for significance. For whole brain analysis of brain glucose metabolism on a voxel-wise basis using the processed FDG PET images, SPM12 (www.fil.ion.ucl.ac.uk/spm/) was used to investigate the effect of genetic variation on brain glucose metabolism across the whole brain. Age and sex were used as covariates. In order to adjust for multiple comparisons, the significant statistical parameters were selected to correspond to a threshold of p-value < 0.05 (FDR-corrected).

### Assessment of SM ratios using targeted metabolomics in ADNI-1

For the investigation of SM ratios measured by targeted metabolomics using the Biocrates P180 kit, we used the same cohort data and statistical models used in Toledo et al.^19^. For selection of the most informative SM ratio, we first calculated all ratios between short-chain (chain length <C20) and long-chain (≥C20) SMs on metabolite levels not adjusted for medication.For each ratio, we then identified significant medications using backward selection based on the Bayesian Information Criterion. Significant medications were included as additional covariates extending the base models described in Toledo et al.^19^ for phenotype associations. Using the P_gain_ criterion, which is defined by the ratio of the minimum association p-value of the constituents of a ratio with the association p-value of the ratio and provides a measure of significance added by the ratio, we obtained the ratio of SM (d34:1) and SM (d43:1) as the one with the largest overall P_gain_.

### Replication analysis of SM ratios using targeted lipidomics in ADNI-1

A more detailed lipidomics method was applied in the ADNI-1 samples to obtain better coverage of the sphingolipidome. Methodology on the ADNI cohort was as previously described^20^. In brief, extracted samples were run using reverse phase liquid chromatography coupled with a triple quadrupole mass spectrometer (Agilent 6490, Agilent). Characterization of sphingolipid isomers have been reported previously^66^ where repeated pooled runs using differing mass spectrometry conditions to obtain structurally informative fragments in MS/MS. Ratios were generated using 112 sphingolipid species and log2-transformed. Linear regression with ADAS-Cog. 13 was done with age, sex, BMI, HDL-C, total cholesterol, clinical triglycerides, fasting status and APOE e4 genotype as covariates. p-values were corrected for multiple correction comparison using the Benjamini and Hochberg approach^67^.

### Candidate mGWAS analysis in ADNI-1

We downloaded genome-wide genotype data for ADNI-1 participants from LONI. Genotype quality control (QC) included exclusion of samples and genotypes with <95% call rate and exclusion of variants that violated a Hardy-Weinberg-Equilibrium (HWE) test p-value of 1 × 10-5 or had a minor allele frequency (MAF) <5%. We then performed autosomal mGWAS analysis with the three SMs (SM (d32:0), SM d(34:1), SM (d38:3)) reported as significantly associated with markers of AD in Toledo et al.^19^, as well as the SM(d43:1)/SM(d34:1) ratio reported here. As covariates, we included age, sex, diagnostic group, as well as the first five components derived by multidimensional scaling (MDS) analysis to account for population stratification. The threshold for genome-wide significance adjusted for four metabolic traits was p-value ≤ 1.25 × 10-8. Genetic associations were calculated using PLINK v1.9^68^.

### Phenotype GWAS and global SM mGWAS analysis in ADNI-1/GO/2

Genome-wide genotyping data of ADNI-1/GO/2 participants were collected using the Illumina Human 610-Quad, HumanOmni Express, and HumanOmni 2.5M BeadChips. Before imputation, standard QC procedures of GWAS data for genetic markers and subjects were performed (variant call rate <95%, HWE test p-value <1 × 10-6, and MAF <1 %, participant call rate <95%, sex check and identity check for related relatives). Then, non-Hispanic Caucasian participants were selected using HapMap 3 genotype data and MDS analysis. Genotype imputation was performed for each genotyping platform separately using the Haplotype Reference Consortium (HRC) reference Panel r1.1 and merged afterwards, resulting in data on 1,576 individuals and 20,779,509 variants. Using this dataset, we ran GWAS analyses for each outcome (A-T-N-C measures, clinical diagnosis and metabolite levels) that included outcome-specific sets of covariates. These are listed in Supplementary Table 6, along with the respective numbers of included individuals and genetic variants.

### Annotation of genetic variants and gene-wide significance thresholds

Previously reported metabolite associations for genes in the SM pathway were retrieved from SNiPA^50^, which was also used to identify overlapping expression quantitative trait loci (eQTLs) from multiple sources. Effect directions of genotype-metabolite and eQTL associations were obtained from the original publications^50^. SNiPA was also used to project genetic variants to genes, a process that includes mapping of variants to genes via genomic location, links to genes via expression and protein QTLs, as well as location in a gene-associated promoter or enhancer region 50. The number of all genetic variants projected to a particular gene was used to derive gene-wise Bonferroni thresholds for significant genetic associations (p-value ≤ 0.05/(number of variants)). Furthermore, as SNPs within genes are correlated due to linkage disequilibrium and Bonferroni correction is often too conservative, we used permutation test, which provides a gene-based empirical p-value that corrects for the number of SNPs within each gene by randomly permuting the phenotypes multi times (20,000 times) and performing statistical tests for all permuted data sets.

### Mouse Model

Experiments were approved by the Division of Comparative Medicine (DCM) from SUNY Downstate Medical Center. APPswe/PS1dE9 (referred to as APP/PS1) and C57Bl/6J (referred to as WT) mice were purchased from The Jackson Laboratory. The APP/PS1 is a double transgenic mouse expressing a chimeric mouse/human amyloid precursor protein (Mo/HuAPP695swe) and a mutant human presenilin 1 (PS1-dE9) both directed to CNS neurons^69^. 50% of animals used in all experiments were males.

### Fingolimod Administration

To determine if fingolimod oral administration achieves appropriate plasma concentration, we treated 8 WT mice at 7 m.o (50% Females) with fingolimod at 1 mg/Kg/day for 4 wks. Plasma samples were collected at two time points (2^nd^ and 4^th^ weeks) after treatment and analyzed by UHPLC and MS-MS. Fingolimod levels in plasma were in ng/ml: 2^nd^ week = 8.03 ± 0.24 and 4^th^ week = 10.02 ± 0.4. The results indicate that oral administration is an appropriate route for mice experiments. We used APP/PS1 and their wildtype littermates to examine fingolimod effects in vivo. Fingolimod treatment was provided in drinking water in a dark container and changed every 48 h to provide 1 mg/Kg/day.

### *In vitro* Electrophysiological recordings

Mice were anesthetized with Ketamine/Xylazine (100/10 mg/kg) and decapitated with an animal guillotine. Horizontal hippocampal slices (400 μm) were prepared using a Vibrotome slicer (VT 1000S; Leica) in ice-cold cutting solution containing the following in mM: 130 potassium gluconate, 5 KCl, 20 HEPES acid, 25 glucose, 0.05 kynurenic acid, 0.05 EGTA-K, and pH equilibrated at 7.4 with KOH. After slicing, the tissue was allowed to recover for an hour before the beginning of experiments in artificial CSF (aCSF) that contained the following in mM: 157 Na^+^, 136 Cl^−^, 2.5 K^+^, 1.6 Mg^2+^, 2 Ca^2+^, 26 HCO_3_^−^, and 11 D-glucose.

LTP recordings were performed in an interface chamber (Fine Scientific Tools, Vancouver Canada) and slices were perfused with aCSF continuously bubbled with 95% O_2_/5% CO_2_, to maintain pH near 7.4 and the temperature was set at 34 °C. Field excitatory post-synaptic potentials (fEPSPs) were recorded in the CA1 stratum radiatum with a glass electrode filled with aCSF (2–3 MΩ resistance). The fEPSPs were elicited by stimulating the Schaffer collateral fibers with a bipolar electrode. Input-output curves were obtained, and a stimulus that evoked ∼40% of the maximum fEPSP was selected to record the baseline. Baseline responses were obtained (15 min with an inter-stimulus interval of 20 s) before high-frequency stimulation (HFS) (one train of 100 stimuli at 100 Hz) was used to induce synaptic LTP. Responses were recorded for 60 min after HFS. The tungsten stimulating electrodes were connected to a stimulus isolation unit (Grass S88), and the recordings were made using an Axoclamp 2B amplifier (Molecular Devices) and then filtered (0.1 Hz to 10 kHz using -6 dB/octave). The voltage signals were digitized and stored on a PC using a DigiData 1200 A (Molecular Devices) for off-line analysis. The fEPSP slope was measured and expressed as a percentage of baseline. The data was analyzed using Axon™ pCLAMP™ software, and the results are expressed as the mean± standard error of the mean (SEM). Data was analyzed statistically using repeated measures ANOVA with the SPSS package.

### Novel Object Recognition (NOR)

Mice were habituated to experimental apparatus consisting of a gray rectangular open field (60 cm x 50 cm x 26 cm) for 5 min in the absence of any objects for 3 days. On the third day, after the habituation trial, mice were placed in the experimental apparatus in the presence of two identical objects and allowed to explore them for 5 min. After a retention interval of 24 h, mice were placed again in the apparatus, where one of the objects was replaced by a novel object. All sessions were recorded using Noldus Media Recorder software. Exploration of the objects was defined as the mice facing and sniffing the objects within 2 cm distance and/or touching them, assessed with ANY-maze software. The ability of the mouse to recognize the novel object (discrimination index) was determined by dividing the mean time exploring the novel object by the mean of the total time exploring the novel and familiar objects during the test session.

### Barnes Maze

The behavioral apparatus consisted of a white flat, circular disk with 20 holes around its perimeter. One hole held the entrance to a darkened escape box not visible from the surface of the board, allowing the subject to exit the maze. The escape chamber position remained fixed during all trials. Mice learn the location of the escape hole using spatial reference points that were fixed in relation to the maze (extra-maze cues). The task consisted of one habituation trial on day 1 where the escape hole was presented to the animal, the animal remained in the escape box for 2 min. After the habituation trial the training phase consisted of four 3-min trials of spatial acquisition for 4 consecutive days with a 15 min inter-trial interval. On the fifth day (probe trial) the escape box was removed, and the animals were allowed to explore the maze for 90 s. All sessions were recorded using Debut video software, and assessed through ANY-maze software. For each trial, several parameters were recorded to assess performance. These include: the latency to locate the escape box, the number of incorrect holes checked prior to entering the escape box, as well as the distance traveled prior to locating the escape box. For the probe trial, time spent in the target quadrant and target hole were analyzed.

## Data Availability

Metabolomics datasets from the AbsoluteIDQ-p180 metabolomics kit used in the current analyses for the ADNI-1 and ADNI-GO/-2 cohorts as well as the RNASeq data from the ROS/MAP, Mount Sinai Brain Bank Cohort, and the Mayo Clinic cohort are available via the Accelerating Medicines Partnership-Alzheimer’s Disease (AMP-AD) Knowledge Portal and can be accessed at http://dx.doi.org/10.7303/syn5592519 (ADNI-1), http://dx.doi.org/10.7303/syn9705278 (ADNI-GO/-2), https://doi.org/10.7303/syn3388564 (ROS/MAP), https://doi.org/10.7303/syn3157743 (MSSB), and https://doi.org/10.7303/syn5550404 (Mayo clinic). The full complement of clinical and demographic data for the ADNI cohorts are hosted on the LONI data sharing platform and can be requested at http://adni.loni.usc.edu/data-samples/access-data/. The full complement of clinical and demographic data for the ROS/MAP cohorts are available via the Rush AD Center Resource Sharing Hub and can be requested at https://www.radc.rush.edu.

## Data Availability

http://dx.doi.org/10.7303/syn9705278

https://doi.org/10.7303/syn3388564

https://doi.org/10.7303/syn3157743

https://doi.org/10.7303/syn5550404

http://adni.loni.usc.edu/data-samples/accessdata/

https://www.radc.rush.edu

## Acknowledgements

Data collection and sharing for this project was funded by the Alzheimer’s Disease Neuroimaging Initiative (ADNI) (National Institutes of Health Grant U01 AG024904) and DOD ADNI (Department of Defense award number W81XWH-12-2-0012). ADNI is funded by the National Institute on Aging, the National Institute of Biomedical Imaging and Bioengineering, and through generous contributions from the following: AbbVie, Alzheimer’s Association; Alzheimer’s Drug Discovery Foundation; Araclon Biotech; BioClinica, Inc.; Biogen; Bristol-Myers Squibb Company; CereSpir, Inc.; Cogstate; Eisai Inc.; Elan Pharmaceuticals, Inc.; Eli Lilly and Company; EuroImmun; F. Hoffmann-La Roche Ltd and its affiliated company Genentech, Inc.; Fujirebio; GE Healthcare; IXICO Ltd.; Janssen Alzheimer Immunotherapy Research & Development, LLC.; Johnson & Johnson Pharmaceutical Research & Development LLC.; Lumosity; Lundbeck; Merck & Co., Inc.; Meso Scale Diagnostics, LLC.; NeuroRx Research; Neurotrack Technologies; Novartis Pharmaceuticals Corporation; Pfizer Inc.; Piramal Imaging; Servier; Takeda Pharmaceutical Company; and Transition Therapeutics. The Canadian Institutes of Health Research is providing funds to support ADNI clinical sites in Canada. Private sector contributions are facilitated by the Foundation for the National Institutes of Health (www.fnih.org). The grantee organization is the Northern California Institute for Research and Education, and the study is coordinated by the Alzheimer’s Therapeutic Research Institute at the University of Southern California. ADNI data are disseminated by the Laboratory for Neuro Imaging at the University of Southern California.

The Religious Orders and the Rush Memory and Aging studies were supported by the National Institute on Aging grants P30AG10161, R01AG15819, R01AG17917, U01AG46152, and U01AG61356. National Institute on Aging (NIA) supported this work (R01 AG059093). NIA also supported the Alzheimer Disease Metabolomics Consortium which is a part of NIA’s national initiatives AMP-AD and M^2^OVE-AD (R01 AG046171, RF1 AG051550, 3U01 AG061359-02S1, and 3U01 AG024904-09S4). Additionally, M.A., R.K.D., and G.K. are supported by NIA grants RF1 AG058942 and R01 AG057452. M.A. and G.K. are also supported by funding from Qatar National Research Fund NPRP8-061-3-011. K.N. is supported by NIH grants NLM R01 LM012535 and NIA R03 AG054936. X.H. is supported by NIA grant RF1 AG061872. P.B. acknowledges the support of 5U01 AG061359-02 and 5U01 AG061359-03 from NIA. C.F. acknowledges the support of R01AG062514, U01AG046139 from NIA.

## Supplementary files

**Supplementary Table 1:** List of enzymes in the SM pathway. The list of enzymes was curated from human genome-scale metabolic reconstruction, Recon 3D.

**Supplementary Table 2:** Differential expression for genes in the SM pathway. The expression changes were analyzed from post-mortem brain samples and compared in AD vs controls.

**Supplementary Table 3:** Genetic variants in the coding regions of genes in SM pathway associated with AD and A-T-N-C measures. Gene-based association analysis pof ADNI individuals were used for identifying the genetic variants.

**Supplementary Table 4:** Sphingolipids profiled in ADNI1 individuals. A total of 112 individual species and 12,544 ratios were identified from the comprehensive lipidomics analysis.

**Supplementary Table 5:** Genome-wide and gene-wide significant associations for 14 related SMs for six genes in the pathway.

**Supplementary Table 6:** GWAS analyses for A-T-N-C measures, clinical diagnosis and metabolites levels in 1,576 individuals

**Supplementary Figure 1:** Association of genetic variants in *CERS3* with CSF t-tau biomarker

**Supplementary Figure 2:** Latency analysis in Fingolimod treated vs untreated APP/PS1 mice. Comparison of APP/PS1 Fingolimod treated vs. untreated APP/PS1 (9 m.o. 50% n=6 treated, 4 untreated), analysis of the latency during training shows a deficit in APP/PS1 untreated compared to APP/PS1 treated s (RM-ANOVA F= 3.1; p = 0.041)

